# The impact of access to financial services on mitigating COVID-19 mortality globally

**DOI:** 10.1101/2022.09.12.22279872

**Authors:** Todd A. Watkins, Khue Nguyen, Hamza Ali, Rishikesh Gummakonda, Jacques Pelman, Brianna Taracena

**Affiliations:** Martindale Center for the Study of Private Enterprise & Department of Economics, Lehigh University, USA; Data for Impact Fellows, Lehigh University, Bethlehem, PA, USA

## Abstract

The COVID-19 pandemic has disproportionately affected different social and demographic groups, deepening the negative health implications of social and economic inequalities and highlighting the importance of social determinants of health. Despite a deep literature on pandemic-related disparities, specifically regarding social determinants and health outcomes, the influence of the accessibility of financial services on health outcomes during COVID-19 remains largely unexplored. Modeling (pre-omicron) COVID-19 mortality across 142 nations, we assess the impact of national-level usage and access to formal financial services. Two financial access indexes constructed through principal component analysis capture (1) usage of and access to formal financial tools and (2) reliance on alternative and informal financial tools. On average, nations with higher pre-pandemic use of and access to formal financial services had substantially lower population mortality risk from COVID-19, controlling for key population health, demographic, and socioeconomic covariates. The scale of effect is similar in magnitude—but opposite in direction—to major risk factors identified in previous literature, such as lung cancer, hypertension, and income inequality. Findings suggest that financial services deserve greater attention both in the public health literature related to COVID-19 and more broadly in policy discussions about fostering better public health overall.

## INTRODUCTION

The COVID-19 pandemic has both highlighted the importance of social determinants of health and exacerbated the negative health implications of social and economic inequalities. There is broad literature and evidence that inequalities in income, access to resources, and other measures of household financial security affect household members’ health [1,2] Specific to the pandemic, two systematic reviews of the burgeoning literature on the social determinants related to COVID-19 health outcomes, one early in the pandemic [3] and one more recently by the World Health Organization [4]—more than 200 articles and reports are included in the latter alone—revealed “glaring inequities” among population groups; main contributing factors include poverty, lack of household resources, affordability of prevention measures, and limited access to various health and social services. Despite the hundreds of such studies related to the pandemic, and the obvious importance of, e.g., ability to pay, affordability, and payment options for accessing health services, none addressed access to broader arrays of financial services beyond formal insurance as a determinant of health outcomes from COVID-19. Yet access to savings, credit, money transfer services, and the like can also serve to buffer risks [5]. The preliminary evidence presented in the sections that follow suggests this may be a major gap in the literature.

Before COVID-19 hit, many developing economies had seen expansions in financial access such as banking deposit and credit services, digital payment systems, and other inclusive fintech innovations. For decades, advocates for global-scale investments in inclusive financial services have championed the role that improved access to financial services plays in helping families in developing economies, particularly those reliant on income from informal work, buffer the risks to their livelihoods and health they face from highly variable and unstable income flows [6–11]. Job losses and economic turmoil due to COVID-19 have exacerbated those risks to an unprecedented degree globally.

Yet it remains an open question whether investments in inclusive financial services and inclusive fintech have mattered for global health. Because health is such a complex socio-economic phenomenon, and financial access is only one (perhaps minor) element among many confounding factors, teasing out the health effects of expanding financial services has proven remarkably challenging. The unfortunately stark and broad natural experiment of COVID-19 might have strengthened the signal-to-noise ratio enough to present a unique opportunity to deepen understanding of the role of access to financial services in health outcomes.

Most of the literature to date linking COVID-19 and financial services has focused on how the pandemic has challenged the financial security of financial sector institutions (e.g., [12–14]), of businesses [15–17], and of households [18–20]. Similarly, the public health literature beyond COVID-19 has focused on how health problems can cause financial risk, including so-called distress financing, the phenomenon of lower-income households coping with health crises and health-related negative income shocks by relying on selling assets and over-indebtedness, in turn begetting further financial risk for the households [21–23]. Distress financing tends to be higher where insurance is thin and national systems lack universal healthcare.

But causality should run the other direction as well if financial security is beneficial to health. Advocates for inclusive finance might have hoped that financial access would have some impact on risk mitigation during the pandemic. Did it? If advocates have been correct, families, communities, and nations that had better financial access pre-COVID-19 should have been better able to weather the health storm in measurable ways during the pandemic. Somewhat surprisingly, there so far has been no global study exploring how the pandemic’s effects correlate with financial access metrics and whether there is any evidence of risk mitigation.

The analysis that follows models COVID-19 mortality rates across 142 nations and finds that nations where residents had greater (pre-pandemic) access to an array of formal financial services had substantially lower population mortality risk during the pandemic. Indeed, the risk reduction is surprisingly large, similar in magnitude to but opposite in direction from the increased COVID-19 mortality risks associated with higher rates of lung cancer, hypertension, and greater income inequality.

## VARIABLES AND DATA SOURCES

Table 1 lists the dependent and control variables, details on the specific metrics used, and data sources; all are publicly available and measured at the national level. The dependent variable is (log-transformed) COVID-19 deaths per million population through the end of September 2021. This cutoff date was before the discovery and spread of the omicron variant. The independent variables listed in Table 1 aim to capture and control for factors known to contribute to variation across nations in COVID-19 mortality rates. All independent variables use the most recently available data predating the pandemic, i.e., 2019 or earlier. Variables with highly skewed distributions are natural log transformed. Two additional variables of main interest are financial services access indexes, constructed as discussed in the next section.

**Table 1.**
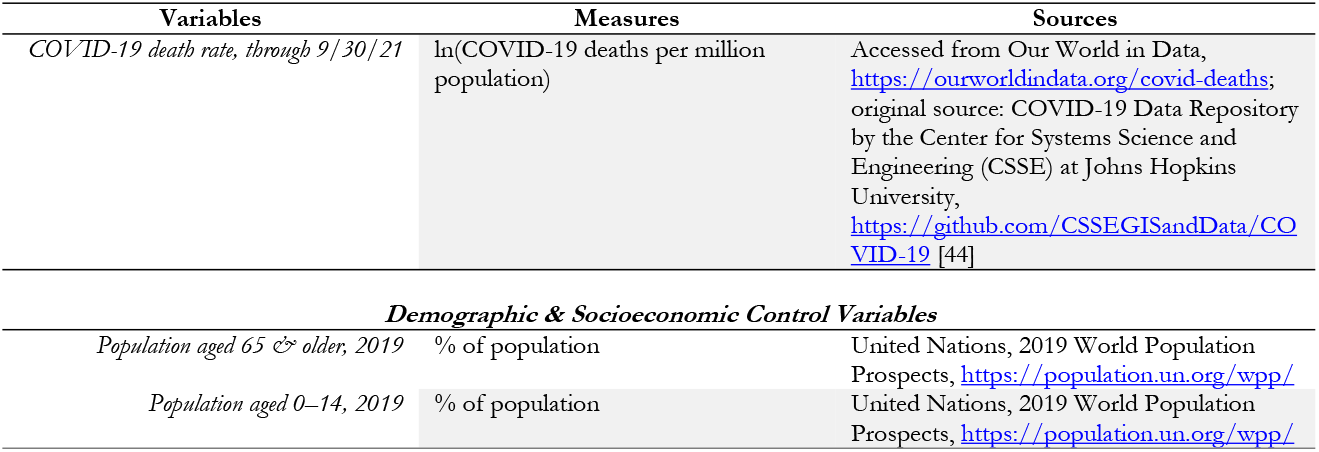
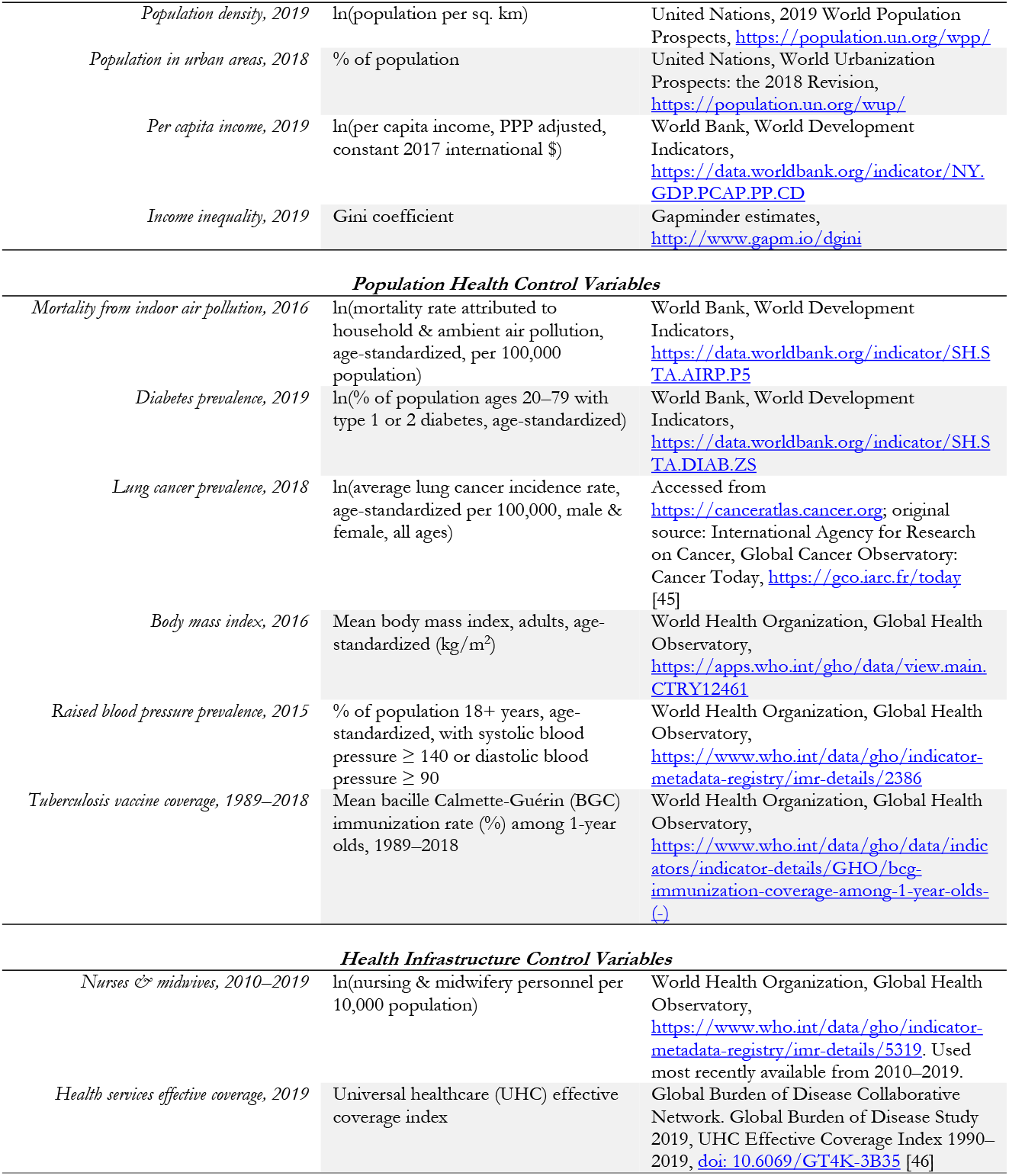
Dependent and Control Variables, with Sources.

Population-level COVID-19 mortality risks are, by now, well understood in the medical literature to be related to a variety of population-level risk factors. Though our main interest here is not these previously studied population health and socioeconomic variables, we include some key national-level public health risk characteristics as control variables. These control variables were selected based on previous evidence in the literature as covariates with COVID-19 mortality, including population health and respiratory issues (levels of hypertension, lung cancer, tuberculosis, diabetes, obesity, and air pollution); healthcare system availability and effectiveness; and socio-economic characteristics, including age distribution, urbanization, population density, and socioeconomic inequalities (e.g., [24–43]).

In addition, we also screened for data availability. Much of the statistical modeling in the national-level literature suffers from limited data availability across nations, restricting sample size. Aiming for the broadest and most representative set of countries, we selected publicly available variables at the national level that had no more than five nations missing from the 142 nations for which financial inclusion variables and COVID-19 mortality rates were also available. Appendix Table 1A shows descriptive statistics for all variables in Table 1, and Table 3A is the correlation matrix.

## FINANCIAL ACCESS VARIABLES METHODOLOGY

To assess the extent to which access to financial services has contributed to mitigating COVID-19 risks, we develop financial inclusion indices at the national level using metrics from the World Bank’s Global Findex Database [47]. The Global Findex includes multiple variables measuring the degree to which the population of each country is engaged in the use of various financial services and activities (as percentage of population). The data are collected through national surveys of more than 150,000 adults from 144 countries, of which 142 also had COVID-19 mortality data available.

We construct our financial access indexes from principal component analysis (PCA) on 20 metrics drawn from the Global Findex Database 2017 panel, the most recent available pre-pandemic. The 2017 panel includes indicators on the access and use of both formal and informal financial services as well as the use of various financial technologies. As described in Table 2, with descriptive statistics in appendix Table 2A, several metrics relate to access to and usage of traditional formal financial tools, such as having any sort of financial institution account, borrowing from and saving in financial institutions, making transactions through financial institutions, and owning debit or credit cards. Other variables measure the use of digital financial services such as making transactions through mobile phones or payments via the internet. We also incorporate several variables indicative of access to financial security tools such as the availability of emergency funds, loans from financial institutions, borrowing for health purposes, and saving for old age.

**Table 2.**
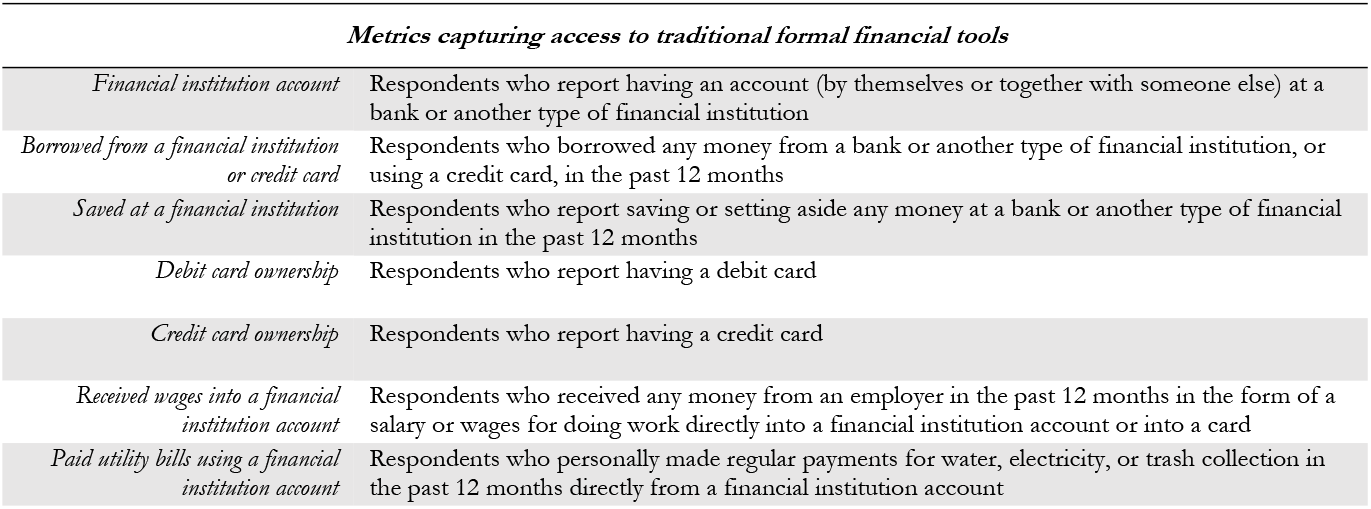
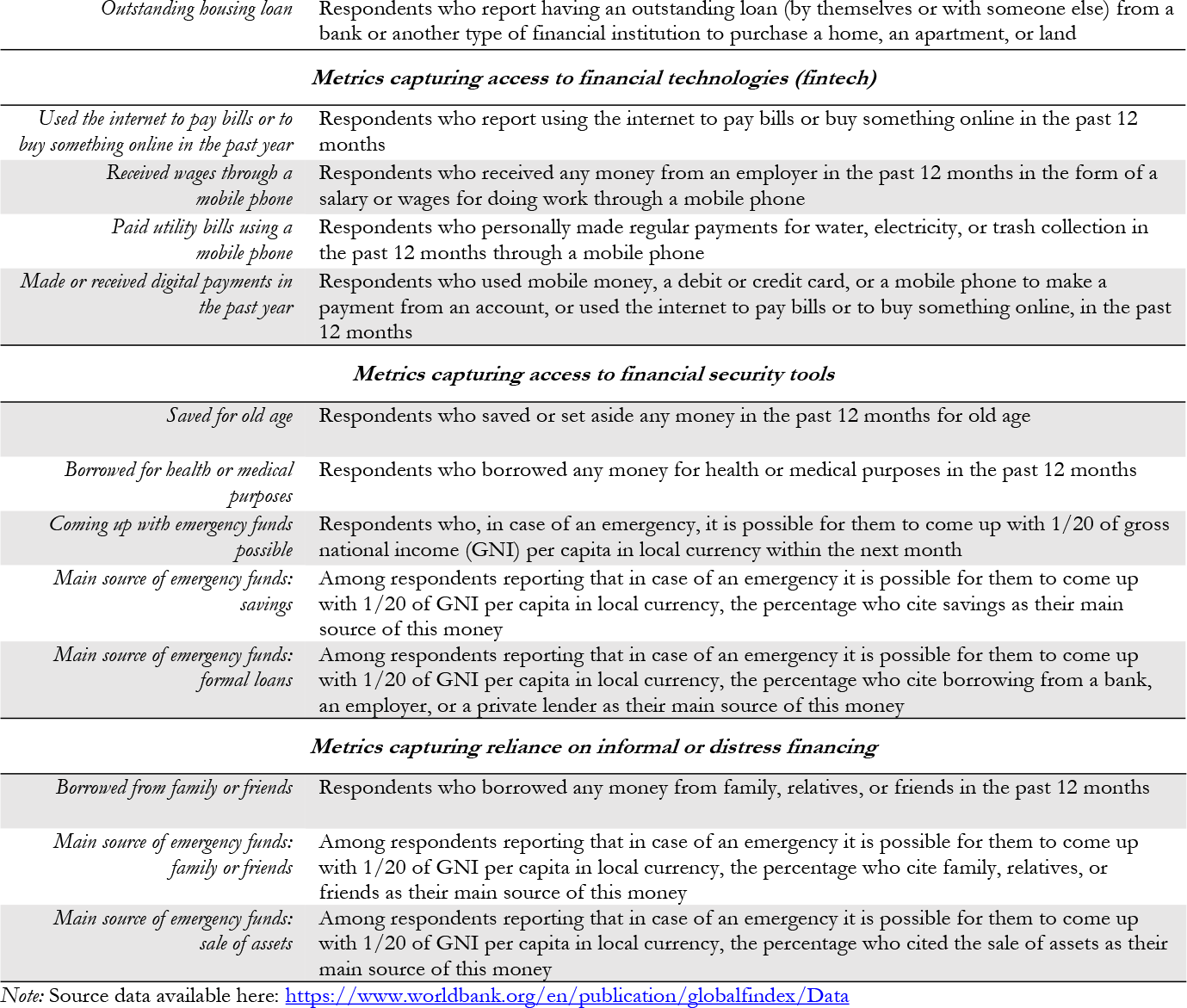
Global Findex Variables Used to Construct Financial Access Indices (% age 15+)

**Table 3.** Identification of Principal Components.

| Component | Eigenvalue | Difference | Proportion of Variance | Cumulative Variance |
| --- | --- | --- | --- | --- |
| *Comp1* | 12.400 | 10.466 | 0.6200 | 0.6200 |
| *Comp2* | 1.935 | 0.630 | 0.0967 | 0.7168 |
| *Comp3* | 1.305 | 0.289 | 0.0653 | 0.7820 |
| *Comp4* | 1.016 | 0.196 | 0.0508 | 0.8329 |
| *Comp5* | 0.821 | 0.351 | 0.0410 | 0.8739 |
| *Comp6* | 0.469 | 0.067 | 0.0235 | 0.8974 |
| *Comp7* | 0.403 | 0.089 | 0.0201 | 0.9175 |
| *etc.* | ... | ... | ... | ... |
| *Comp19* | 0.024 | 0.001 | 0.0012 | 0.9989 |
| *Comp20* | 0.023 | . | 0.0011 | 1.0000 |

Additional variables measure the use of informal or distress finance, e.g., relying on friends and family for borrowing or for emergencies, or selling assets for emergency funding.

To reduce the dimensionality of our exploration of whether countries with greater access to financial services have better success mitigating COVID-19 risks while retaining most of the variability of the underlying metrics, we apply PCA. Our PCA incorporates all 20 variables in Table 2, from the Global Findex Database, to create indices of financial access at the national level. As shown in Table 3, the two principal components with the highest eigenvalues together account for nearly three-quarters (72%) of the total variance among the 20 variables. We use these top two to construct our two financial access indexes.

Table 4 shows how the top components correlate with each original variable. The first component correlates positively and relatively evenly across most of the formalized financial tools including both traditional tools (e.g., savings, loans, cards, and other services provided by financial institutions) and using the internet, and it correlates negatively with informal and distress finance, such as relying on family and friends and borrowing for health purposes or selling assets for emergency funds. Thus, we interpret the first component as an index of the extent to which a nation exhibits “Broad access to and use of formal financial tools.” The higher the index, the more widespread and diverse are the formal financial tools in regular use in that country.

**Table 4.** Principal Components’ Correlation with Original Global Findex Variables.

| Variable | Comp1 | Comp2 | Comp3 | Comp4 |
| --- | --- | --- | --- | --- |
| *Finance institution account* | 0.259 | -0.030 | 0.197 | -0.027 |
| *Borrowed from a financial institution or credit card* | 0.261 | 0.009 | -0.012 | 0.155 |
| *Saved at a financial institution* | 0.268 | 0.058 | -0.137 | -0.051 |
| *Debit card ownership* | 0.264 | -0.008 | 0.153 | -0.056 |
| *Credit card ownership* | 0.263 | -0.017 | -0.069 | 0.098 |
| *Received wages into financial institution* | 0.267 | -0.012 | 0.130 | -0.011 |
| *Paid utility bills using financial institution account* | 0.267 | 0.012 | 0.006 | 0.053 |
| *Outstanding housing loan* | 0.248 | 0.080 | 0.030 | 0.032 |
| *Used the internet for online transaction* | 0.272 | 0.066 | 0.032 | -0.021 |
| *Received wages through mobile phone* | 0.061 | 0.535 | 0.232 | -0.259 |
| *Paid utility bills through mobile phone* | 0.179 | 0.427 | 0.081 | -0.125 |
| *Made or received digital payments* | 0.264 | 0.086 | 0.170 | -0.035 |
| *Saved for old age* | 0.255 | 0.002 | -0.190 | -0.030 |
| *Borrowed for health or medical purposes* | -0.193 | 0.312 | 0.003 | 0.023 |
| *Coming up with emergency funds: possible* | 0.194 | -0.234 | 0.014 | -0.167 |
| *Main source of emergency funds: savings* | 0.246 | -0.020 | -0.275 | -0.142 |
| *Main source of emergency funds: formal loan* | 0.077 | 0.173 | 0.269 | 0.850 |
| *Borrowed from family or friends* | -0.163 | 0.368 | 0.251 | -0.237 |
| *Main source of emergency funds: family/ friends* | -0.134 | -0.247 | 0.620 | -0.055 |
| *Main source of emergency funds: sale of assets* | -0.138 | 0.361 | -0.419 | 0.202 |

By contrast, the second index associates most closely with metrics related to alternative financial tools, such as mobile phone transactions and informal finance of borrowing from family and friends, and distress financing tools, such as selling assets for emergency uses and borrowing for health and medical needs. It is also negatively associated with the ability to raise emergency funds and access to family funding for emergencies, suggesting that high values of the second index associate, in part, with financial resilience challenges. We therefore interpret the second component as an index capturing a nation’s tendency toward “Reliance on alternative, informal, and distress financial tools.” Descriptive statistics on two financial tools indexes created from these two principal components are shown in appendix Table 1A. The influence of these two index variables on COVID-19 mortality is our main interest in the econometric models that follow.

## MODELS AND RESULTS

As the slight differences among variable observation counts in appendix Table 1A indicate, ten nations lack values for one or several of the control variables, but the specific missing variables differed among nations. Because those ten nations did have data for most of our other variables, rather than lose them from the analysis, we opted for a linear model using full information maximum likelihood (FIML) estimation (following [48], which shows FIML estimates are similar to those from established missing variable imputation methods, e.g., [49–52]. Because COVID-19 patterns correlate geographically, we include World Bank region dummies, and robust standard errors are adjusted clustered by region. Our main results are shown in Table 5 (repeated as Model 1 in Table 4A), followed by a comparison OLS model in Table 6 without those ten nations that lacked full data (Model 2 in Table 4A), again with robust standard errors adjusted clustered by region, as a robustness check. Either model explains more than 72% of the variation in COVID-19 mortality rates across the 142 nations.

**Table 5.**
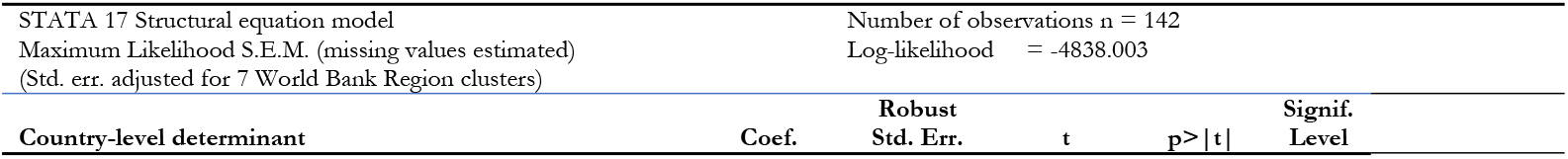
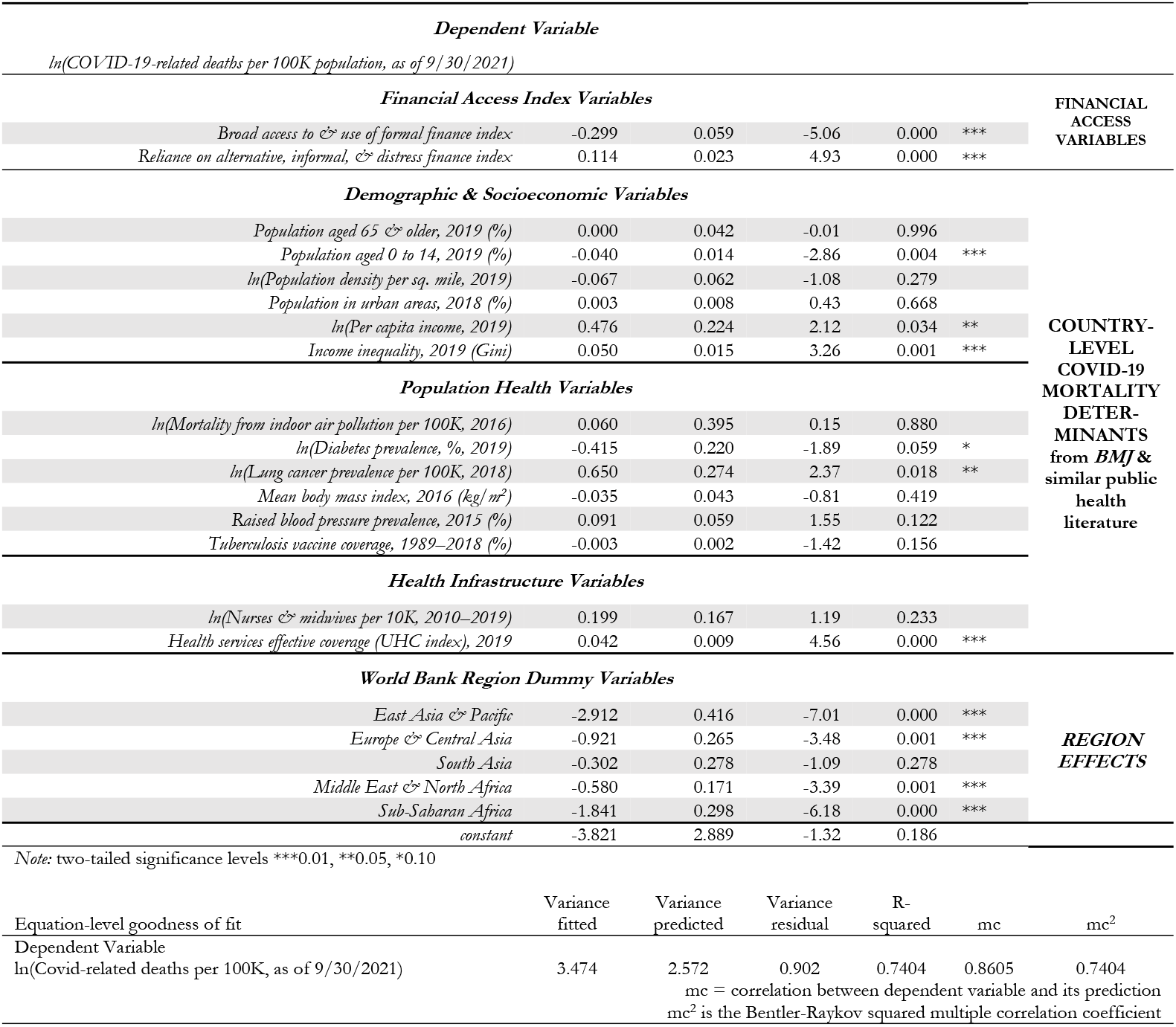
Full Information Maximum Likelihood Estimation.

**Table 6.**
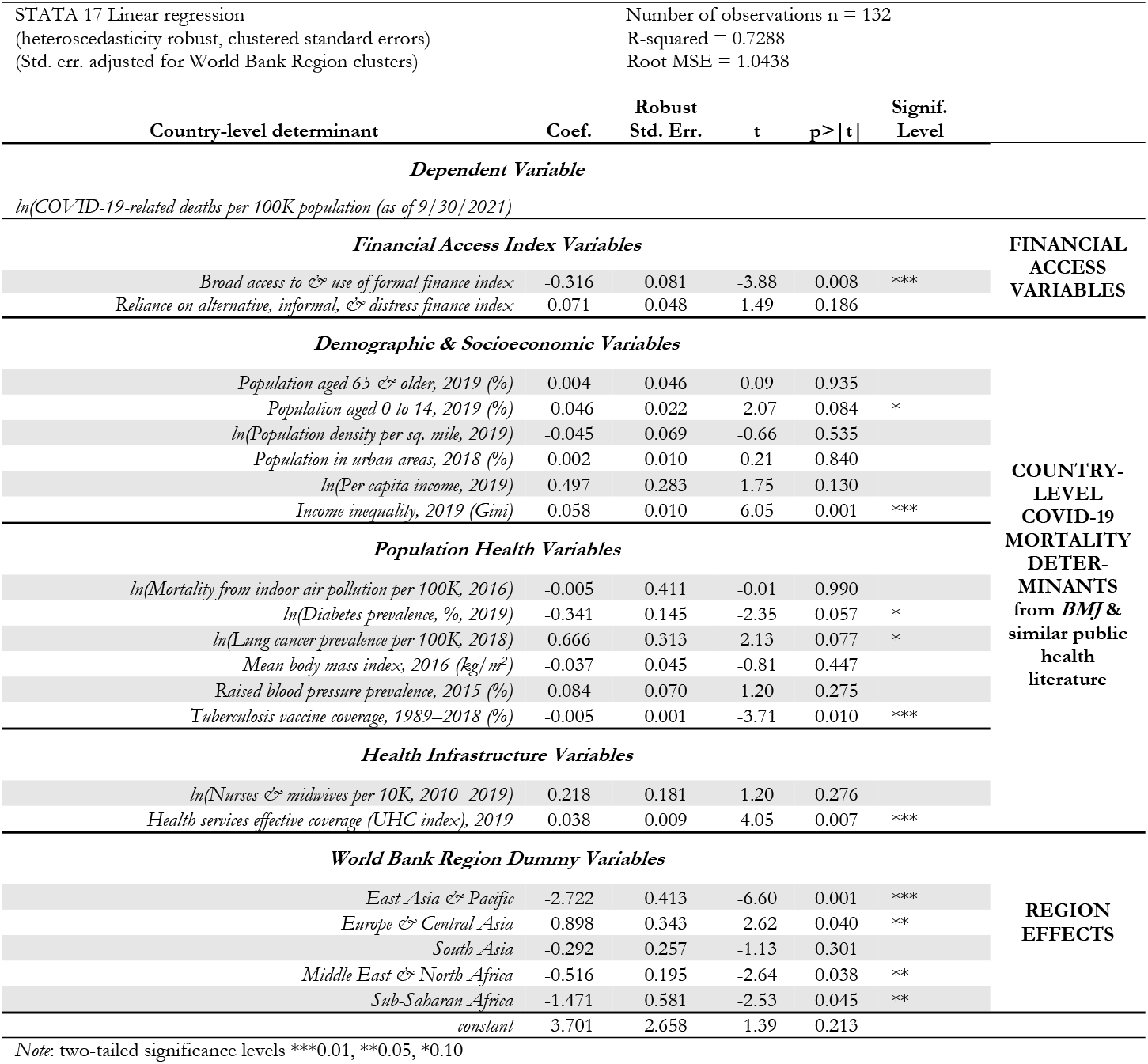
OLS Regression Using Only Nations with Complete Information.

The first financial access index, capturing access to formal financial services, is statistically significantly related to COVID-19 mortality rates (p<.001), controlling for demographic, socioeconomic, population health and health infrastructure variables, and region. In Table 5, the −0.299 coefficient suggests that (pre-omicron) COVID-19 mortality rises 29.9% for a one-unit reduction in the (pre-COVID-19) access to formal finance index. The standard deviation of the index is 3.521, implying that, on average, nations with one standard deviation lower access to formal financial services had COVID-19 mortality rates more than double (0.299*3.521=105%) nations with average financial access, conditional on the other COVID-19 mortality covariates. The alternative coefficient estimate (0.316) from Table 6 is also statistically significant (p=.008) and very similar in scale, again suggesting more than doubling the mortality rate (111%).

As additional checks on the stability of estimates of our main financial access index variables of interest, appendix Table 4A expands the number of alternative model specifications, showing models with only various subsets of the independent variables. Models 1 and 2 repeat Tables 5 and 6. Model 3 removes the region dummies. Model 4 uses only the two financial tools indexes. Because financial system strength is highly correlated with national income, Model 5 adds only a single other variable, per capita income, to the two financial indexes. Model 6 adds only the other demographic and socioeconomic variables. Model 7 includes only the health infrastructure variables with the financial indexes. Model 8 adds population health variables.

Model 9 aims for parsimony by removing from Model 2, post OLS, any variable with p>0.4. (Not shown, but the variables included in Model 9 are the same as those identified by a different technique, LASSO regression, except the LASSO-suggested model includes population density, which when added has essentially no substantive effect on other significant coefficients.) Finally, Model 10 includes the same variables as Model 9 but employs the FIML estimation method (as used in Model 1) to enable including the ten nations missing one or more specific variables. A Ramsey RESET test and a LINK test are conducted after each model (except the FIML models) to test functional form and omitted variable problems. The only statistically significant evidence of problems arises in the Models (4, 5, and 7) that omit all or most control variables.

Appendix Table 4A shows the estimated size of the relationship between COVID-19 mortality rate— approximate doubling per standard deviation reduction in the index of access to formal financial services— robust to alternative sets of control variables, except in Model 4, which only includes the two financial tools indexes. Compared to Model 4, the addition in Model 5 of a single additional covariate that is highly positively correlated with the depth of financial systems, per capita income, switches the sign of the coefficient on the first financial index. A similar effect can be seen in Model 7, which adds only the two health infrastructure variables, both also strongly positively correlated with national wealth. In other words, access to formal financial services becomes significantly negatively associated with COVID-19 mortality once any variables also related to national wealth are controlled for.

By contrast, the results for the second financial services index, reliance on alternative, informal, and distress financing, are weaker in scale and statistically less robust. In Table 5, a nation’s pre-COVID-19 tendency to rely on alternative, informal, and distress financial tools is positively associated with higher COVID-19 mortality and statistically significant (p<.001). According to this model, mitigating COVID-19 impacts apparently was more challenging where financial emergency coping mechanisms rely on informal options and distress financing—an intuitively appealing result. However, compared to the formal finance index, the strength of the association is weaker. A one standard deviation increase in the alternative, informal, and distress finance index is associated with a 15.9% (1.391*.114) increase in mortality. In the OLS version, Table 6, which includes data from fewer nations, the second index, though still positively related to COVID-19 mortality, falls to statistical insignificance (p=0.186). Moreover, as appendix Table 4A shows, coefficient estimates and significance for this second index are sensitive to which control variables are included.

While the control variables—drawn from the medical and public health literatures—are not our central interest, we now turn to those population health, demographic, and socioeconomic variables. Youthful populations are associated with statistically significantly lower mortality, as are more-equal income distributions. Consistent with existing literature (e.g., [53–56]), nations with higher degrees of income inequality have statistically significantly higher COVID-19 mortality rates. A one-unit increase in national Gini coefficient raises expected COVID-19 mortality rate by about 5%–6%, depending on the model. Nations with income inequality one standard deviation (7.79) above the average level would have roughly 40%–50% higher mortality rates.

Given that COVID-19 attacks the respiratory-system [35,36,43], it is unsurprising that population respiratory health appears important too. Higher national pre-COVID-19 lung cancer prevalence is a statistically significant COVID-19 mortality risk factor in models with regional dummies. As shown in Figure 1.b, a one-unit increase (approximately the standard deviation, 1.08) from the mean ln(lung cancer prevalence) relates to a two-thirds greater COVID-19 mortality, conditional on the other independent variables. So too, higher pre-COVID-19 childhood tuberculosis vaccination rates are negatively correlated with COVID-19 mortality (though the latter is not statistically significant in Models 1 and 8).

**Fig 1.**
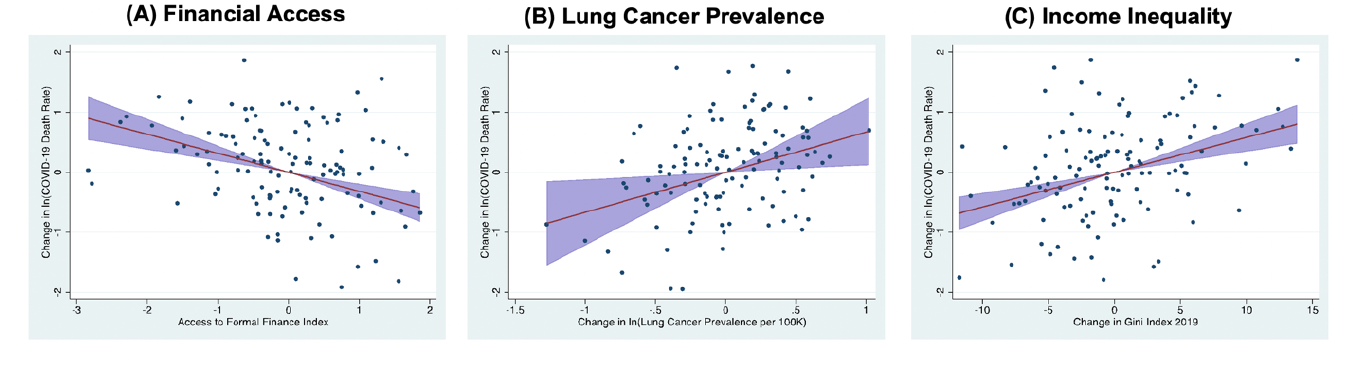
Comparing conditional partial correlation of COVID-19 mortality with financial access, lung cancer, and income inequality. (A) Conditional partial correlation of national COVID-19 mortality with pre-COVID-19 Index of access to formal finance; 95% confidence interval shaded. (B) Similar, with pre-COVID-19 ln(average lung cancer incidence rate, age-standardized per 100,000, male & female, all ages). (C) Similar, with pre-COVID Gini index of income inequality.

We also find that nations with higher per capita incomes had statistically significantly higher mortality rates, even after controlling for characteristics associated with wealth, like aging and obesity, that might be problematic for richer nations. Though curious, this finding is consistent with most of the COVID-19 literature. For example, Goldberg and Reed [28] found, after controlling for several demographic, public health, policy response, weather, and mobility variables, that for a 1% increase in per capita GDP, COVID-19 mortality rose by 0.9%. Our results suggest a smaller relation, roughly half that, but still positive. Reasons for this counteractive yet widely duplicated result are not well understood and have been discussed at length elsewhere (e.g., [28,37–39,56]).

Similarly counterintuitive, nations with more effective healthcare systems, as measured by the universal healthcare effective coverage index, have significantly higher COVID-19 mortality. The size of the relationship is similar to that of lung cancer relation: a one standard deviation increase in the UHC index goes with an approximate two-thirds increase in mortality. Greater population shares of nurses and midwifery personnel are also positively related to mortality, however not statistically significantly in most models. The coefficients on diabetes hint at another puzzle. In models with regional dummies, diabetes prevalence associates negatively with COVID-19 mortality—not the direction we would expect based on the medical literature [41,42]. However, the statistical significance is marginal (p>.05) except in Model 9. Indoor air pollution, body mass index, and raised blood pressure prevalence are not statistically related to mortality in most models.

## DISCUSSION AND CONCLUSIONS

In summary, we find that greater pre-pandemic national levels of use and access to formal financial services are related to substantially lower death rates from (pre-omicron) COVID-19. The result suggests that financial services deserve substantially greater attention both in the public health literature related to COVID-19 and more broadly in policy discussions about fostering better public health overall. Robust financial services like savings, insurance, credit, payment systems, and the like are clearly potentially useful tools for households paying for food, housing, medicines, and health services and dealing with medical emergencies. Yet despite the extensive literature linking household financial security and health and the obvious potential link to COVID-19 outcomes, we are unaware of any other study exploring how COVID-19 mortality relates to financial systems.

To help assess the relative importance of the financial access variable, Figure 1.A, based on the model in Table 5, shows the partial correlation of COVID-19 mortality with pre-COVID-19 levels of access to formal financial services conditional on the other independent variables in Table 5, with 95% confidence interval. The strength of the association in Figure 1.a of COVID-19 mortality with pre-COVID-19 levels of access to formal financial services is similar in scale but opposite in direction to the estimated relations with pre-COVID-19 lung cancer rates (Figure 1.B) and with income inequality (Figure 1.C)—both of which are among the most important determinants of COVID-19 mortality in the medical and public health literatures. Many dozens (perhaps hundreds) of pandemic-related studies have now looked at each of those variables; financial services have been essentially ignored. Considering that the scale of the relationship to population-level COVID-19 mortality rivals that of lung cancer, this appears to be a major exploratory blind spot.

Though the GDP-COVID-19 puzzle is not our central interest, our finding that deeper formal financial systems appear associated with better risk mitigation if anything deepens the perplexity. It is puzzling too that not only the income variable but also both health infrastructure variables relate positively to COVID-19 mortality. All three behave oppositely to our formal financial access metric, despite the clear relationship between national wealth and the depths of both financial systems and health systems. Some analysts have speculated the positive association of national income and COVID-19 might be an artifact of nations with deeper healthcare systems better tracking health statistics [39]. Yet controlling for the healthcare infrastructure here does not solve the GDP riddle. Other speculations include previous experience in lower-income nations with handling and/or accumulated immunities from similar viral (e.g., SARS) outbreaks. Regardless, the jury is still out on why rich countries have suffered more. It is remarkably counterintuitive that that pattern is replicated in both income and healthcare infrastructure.

Untangling the complex socio-economic and socio-structural determinants of health inequalities remains a major challenge. The role of financial tools in health should feature centrally in that inquiry.

## Data Availability

All data used in the study is from publicly available sources, as comprehensively listed in the manuscript.

https://www.worldbank.org/en/publication/globalfindex/Data

https://doi.org/10.6069/GT4K-3B35

https://www.who.int/data/gho/indicator-metadata-registry/imr-details/5319

https://www.who.int/data/gho/data/indicators/indicator-details/GHO/bcg-immunization-coverage-among-1-year-olds-(-)

https://www.who.int/data/gho/indicator-metadata-registry/imr-details/2386

https://apps.who.int/gho/data/view.main.CTRY12461

https://canceratlas.cancer.org

https://gco.iarc.fr/today

https://data.worldbank.org/indicator/SH.STA.DIAB.ZS

https://data.worldbank.org/indicator/SH.STA.AIRP.P5

### APPENDIX

**Table 1A.** Descriptive Statistics for Model Variables.

| Variable | Obs. | Mean | Std. Dev. | Min. | Max. |
| --- | --- | --- | --- | --- | --- |
| *Dependent Variable* |  |  |  |  |  |
| *ln(COVID-19 death rate per million, to 9/30/21)* | 142 | 5.77 | 1.87 | -0.21 | 8.7 |
| *Demographic & Socioeconomic Variables* |  |  |  |  |  |
| *Population aged 65 & older, 2019 (%)* | 142 | 10.11 | 6.91 | 1.26 | 28.4 |
| *Population aged 0–14, 2019 (%)* | 142 | 26.11 | 10.39 | 12.3 | 49.5 |
| *ln(Population density per sq. mile, 2019)* | 142 | 4.34 | 1.40 | 0.75 | 9.03 |
| *Population in urban areas, 2018 (%)* | 139 | 61.81 | 22.13 | 16 | 100 |
| *ln(Per capita income, 2019)* | 141 | 9.50 | 1.17 | 6.72 | 11.7 |
| *Income inequality, 2019 (Gini)* | 142 | 38.08 | 7.79 | 25 | 64 |
| *Population Health Variables* |  |  |  |  |  |
| *ln(Mortality from indoor air pollution per 100K, 2016)* | 139 | 4.05 | 0.99 | 1.95 | 5.78 |
| *ln(Diabetes prevalence, %, 2019)* | 138 | 1.85 | 0.49 | 0 | 2.99 |
| *ln(Lung cancer prevalence per 100K, 2018)* | 139 | 2.40 | 1.08 | -0.77 | 4.08 |
| *Mean body mass index, 2016 (kg/m<sup>2</sup>)* | 139 | 25.50 | 2.03 | 20.6 | 29.6 |
| *Raised blood pressure prevalence, 2015 (%)* | 140 | 24.35 | 4.83 | 11 | 33.4 |
| *Tuberculosis vaccine coverage, 1989–2018 (%)* | 142 | 61.89 | 31.31 | 0 | 99 |
| *Health Infrastructure Variables* |  |  |  |  |  |
| *ln(Nurses and midwives per 10K, 2010–2019)* | 139 | 3.28 | 1.24 | -2.85 | 5.38 |
| *Health services effective coverage (UHC index), 2019* | 141 | 63.79 | 16.42 | 22.3 | 96.34 |
| *World Bank Region Dummy Variables* |  |  |  |  |  |
| *East Asia & Pacific* | 142 | 0.12 | 0.33 | 0 | 1 |
| *Europe & Central Asia* | 142 | 0.32 | 0.47 | 0 | 1 |
| *South Asia* | 142 | 0.04 | 0.20 | 0 | 1 |
| *Middle East & North Africa* | 142 | 0.11 | 0.32 | 0 | 1 |
| *Sub-Saharan Africa* | 142 | 0.25 | 0.43 | 0 | 1 |
| *Financial Access Indexes from Principal Components Analysis* |  |  |  |  |  |
| *Broad access to and use of formal finance index* | 142 | 0 | 3.521 | -4.858 | 9.056 |
| *Reliance on alternative, informal,44 & distress finance index* | 142 | 0 | 1.391 | -2.342 | 4.978 |

**Table 2A.** Summary Descriptive Statistics, Global Findex Variables.

| Variable (% of population aged 15+) | Obs. | Mean | Std. Dev. | Min. | Max. |
| --- | --- | --- | --- | --- | --- |
| *Finance institution account* | 142 | 0.585 | 0.294 | 0.086 | 0.999 |
| *Borrowed from a financial institution or credit card* | 142 | 0.240 | 0.187 | 0.024 | 0.828 |
| *Saved at a financial institution* | 142 | 0.238 | 0.197 | 0.016 | 0.793 |
| *Debit card ownership* | 142 | 0.443 | 0.314 | 0.017 | 0.988 |
| *Credit card ownership* | 142 | 0.193 | 0.210 | 0.001 | 0.826 |
| *Received wages into financial institution* | 142 | 0.246 | 0.199 | 0.016 | 0.683 |
| *Paid utility bills using financial institution account* | 142 | 0.231 | 0.240 | 0.000 | 0.813 |
| *Outstanding housing loan* | 142 | 0.130 | 0.108 | 0.011 | 0.543 |
| *Used the internet for online transaction* | 142 | 0.278 | 0.262 | 0.005 | 0.895 |
| *Received wages through mobile phone* | 142 | 0.025 | 0.028 | 0.000 | 0.134 |
| *Paid utility bills through mobile phone* | 142 | 0.068 | 0.074 | 0.000 | 0.371 |
| *Made or received digital payments* | 142 | 0.541 | 0.283 | 0.073 | 0.994 |
| *Saved for old age* | 142 | 0.210 | 0.155 | 0.024 | 0.614 |
| *Borrowed for health or medical purposes* | 142 | 0.100 | 0.066 | 0.006 | 0.311 |
| *Coming up with emergency funds: possible* | 142 | 0.554 | 0.165 | 0.165 | 0.927 |
| *Main source of emergency funds: savings* | 142 | 0.296 | 0.197 | 0.026 | 0.875 |
| *Main source of emergency funds: formal loan* | 142 | 0.059 | 0.044 | 0.002 | 0.243 |
| *Borrowed from family or friends* | 142 | 0.229 | 0.107 | 0.037 | 0.526 |
| *Main source of emergency funds: family/friends* | 142 | 0.317 | 0.155 | 0.052 | 0.711 |
| *Main source of emergency funds: sale of assets* | 142 | 0.042 | 0.055 | 0.000 | 0.281 |

**Table 3A.** Correlation Matrix.

| Variable | *ln(COVID-19 death rate per million, to 9/30/21)* | *Broad access to & use of formal finance index* | *Reliance on alternative, informal, & distress finance index* | *Population aged 65 & older, 2019 (%)* | *Population aged 0–14, 2019 (%)* | *ln(Population density per sq. mile, 2019)* | *Population in urban areas, 2018 (%)* | *ln(Per capita income, 2019)* | *Income inequality, 2019 (Gini)* | *ln(Mortality from indoor air pollution per 100K, 2016)* | *ln(Diabetes prevalence, %, 2019)* | *ln(Lung cancer prevalence per 100K, 2018)* | *Mean body mass index, 2016 (kg/m<sup>2</sup>)* | *Raised blood pressure prevalence, 2015 (%)* | *Tuberculosis vaccine coverage, 1989–2018 (%)* | *ln(Nurses & midwives per 10K, 2010–2019)* | *Health services effective coverage (UHC index), 2019* |
| --- | --- | --- | --- | --- | --- | --- | --- | --- | --- | --- | --- | --- | --- | --- | --- | --- | --- |
| *ln(COVID-19 death rate per million, to 9/30/21)* | 1 |  |  |  |  |  |  |  |  |  |  |  |  |  |  |  |  |
| *Broad access to & use of formal finance index* | 0.31 | 1 |  |  |  |  |  |  |  |  |  |  |  |  |  |  |  |
| *Reliance on alternative, informal, & distress finance index* | -0.31 | -0.01 | 1 |  |  |  |  |  |  |  |  |  |  |  |  |  |  |
| *Population aged 65 & older, 2019 (%)* | 0.50 | 0.78 | -0.34 | 1 |  |  |  |  |  |  |  |  |  |  |  |  |  |
| *Population aged 0–14, 2019 (%)* | -0.60 | -0.75 | 0.40 | -0.85 | 1 |  |  |  |  |  |  |  |  |  |  |  |  |
| *ln(Population density per sq. mile, 2019)* | -0.02 | 0.18 | -0.26 | 0.18 | -0.28 | 1 |  |  |  |  |  |  |  |  |  |  |  |
| *Population in urban areas, 2018 (%)* | 0.45 | 0.64 | -0.19 | 0.49 | -0.61 | 0.02 | 1 |  |  |  |  |  |  |  |  |  |  |
| *ln(Per capita income, 2019)* | 0.58 | 0.84 | -0.23 | 0.75 | -0.88 | 0.17 | 0.77 | 1 |  |  |  |  |  |  |  |  |  |
| *Income inequality, 2019 (Gini)* | -0.15 | -0.38 | 0.29 | -0.44 | 0.44 | -0.15 | -0.14 | -0.38 | 1 |  |  |  |  |  |  |  |  |
| *ln(Mortality from indoor air pollution per 100K, 2016)* | -0.50 | -0.87 | 0.17 | -0.78 | 0.77 | -0.08 | -0.70 | -0.83 | 0.30 | 1 |  |  |  |  |  |  |  |
| *ln(Diabetes prevalence, %, 2019)* | 0.33 | 0.10 | -0.27 | 0.08 | -0.37 | 0.09 | 0.28 | 0.34 | -0.08 | -0.23 | 1 |  |  |  |  |  |  |
| *ln(Lung cancer prevalence per 100K, 2018)* | 0.62 | 0.64 | -0.38 | 0.78 | -0.86 | 0.14 | 0.52 | 0.78 | -0.48 | -0.68 | 0.35 | 1 |  |  |  |  |  |
| *Mean body mass index, 2016 (kg/m<sup>2</sup>)* | 0.60 | 0.38 | -0.25 | 0.38 | -0.55 | -0.13 | 0.67 | 0.64 | -0.21 | -0.55 | 0.48 | 0.53 | 1 |  |  |  |  |
| *Raised blood pressure prevalence, 2015 (%)* | -0.21 | -0.58 | 0.08 | -0.33 | 0.47 | -0.22 | -0.64 | -0.58 | 0.03 | 0.68 | -0.27 | -0.37 | -0.38 | 1 |  |  |  |
| *Tuberculosis vaccine coverage, 1989–2018 (%)* | -0.04 | -0.58 | 0.02 | -0.41 | 0.29 | -0.20 | -0.29 | -0.35 | 0.30 | 0.44 | 0.02 | -0.28 | -0.13 | 0.29 | 1 |  |  |
| *ln(Nurses & midwives per 10K, 2010–2019)* | 0.55 | 0.65 | -0.21 | 0.64 | -0.73 | 0.07 | 0.53 | 0.75 | -0.41 | -0.64 | 0.23 | 0.70 | 0.53 | -0.36 | -0.24 | 1 |  |
| *Health services effective coverage (UHC index), 2019* | 0.51 | 0.88 | -0.17 | 0.79 | -0.80 | 0.19 | 0.69 | 0.87 | -0.38 | -0.90 | 0.20 | 0.71 | 0.49 | -0.63 | -0.43 | 0.67 | 1 |

**Table 4A.** Alternative Model Specifications.

| Dependent variable<br>ln(COVID-19 death rate per million through<br>9-30-2021) | (1)<br>FIML-SEM<br>All Vars | (2)<br>OLS<br>All Vars | (3)<br>No Region<br>Dummies | (4)<br>Fin Indexes<br>Only | (5)<br>Fin Indexes<br>w/ Income | (6)<br>Demogr &<br>Socio-Econ | (7)<br>Health<br>Infrastr. | (8)<br>Pop.<br>Health &<br>Infrastr. | (9)<br>Post OLS<br>remove<br>p>.4 | (10)<br>FIML-SEM<br>Model 9<br>Vars |
| --- | --- | --- | --- | --- | --- | --- | --- | --- | --- | --- |
| **Financial Access Index Variables** |  |  |  |  |  |  |  |  |  |  |
| *Broad access to & use*<br>*of formal finance index* | -0.2990***<br>(0.0591) | -0.3160***<br>(0.0814) | -0.4099**<br>(0.1212) | 0.1609<br>(0.1143) | -0.2655*<br>(0.1138) | -0.3631**<br>(0.1334) | -0.2934***<br>(0.0725) | -0.3507**<br>(0.1230) | -0.3094***<br>(0.0639) | -0.2993***<br>(0.0548) |
| *Reliance on alternative, informal,*<br>*& distress finance index* | 0.1142***<br>(0.0232) | 0.0714<br>(0.0478) | 0.0107<br>(0.0628) | -0.4117**<br>(0.1147) | -0.1351<br>(0.1135) | 0.0141<br>(0.0888) | -0.1624*<br>(0.0756) | 0.0223<br>(0.0739) | 0.0829*<br>(0.0345) | 0.1234***<br>(0.0290) |
| **Demographic & Socioeconomic Variables** |  |  |  |  |  |  |  |  |  |  |
| *Population aged 65 & older (%)* | -0.0002<br>(0.0416) | 0.0039<br>(0.0457) | 0.0181<br>(0.0397) |  |  | 0.0865**<br>(0.0278) |  |  |  |  |
| *Population aged 0–14 (%)* | -0.0405***<br>(0.0141) | -0.0456*<br>(0.0221) | -0.0551**<br>(0.0201) |  |  | -0.0659<br>(0.0347) |  |  | -0.0466**<br>(0.0153) | -0.0322**<br>(0.0152) |
| *ln(Population density per sq. mile)* | -0.0669<br>(0.0618) | -0.0452<br>(0.0687) | 0.0152<br>(0.0892) |  |  | -0.1812<br>(0.1136) |  |  |  |  |
| *Population in urban areas (%)* | 0.0034<br>(0.0078) | 0.0022<br>(0.0104) | 0.0038<br>(0.0092) |  |  | 0.0068<br>(0.0091) |  |  |  |  |
| *ln(Per capita income)* | 0.4763**<br>(0.2242) | 0.4968<br>(0.2832) | 0.3080<br>(0.2346) |  | 1.5492***<br>(0.2385) | 0.9354***<br>(0.2219) |  |  | 0.5397**<br>(0.1617) | 0.5002***<br>(0.1587) |
| *Income inequality (Gini)* | 0.0501***<br>(0.0154) | 0.0579***<br>(0.0096) | 0.0620***<br>(0.0084) |  |  | 0.0238<br>(0.0289) |  |  | 0.0578***<br>(0.0092) | 0.0499***<br>(0.0168) |
| **Population Health Variables** |  |  |  |  |  |  |  |  |  |  |
| *ln(Mortality from indoor air*<br>*pollution per 100K)* | 0.0596<br>(0.3955) | -0.0054<br>(0.4110) | -0.5089<br>(0.4413) |  |  |  |  | -0.6715<br>(0.3622) |  |  |
| *ln(Diabetes prevalence, %)* | -0.4154*<br>(0.2200) | -0.3411*<br>(0.1454) | 0.0013<br>(0.3167) |  |  |  |  | 0.1759<br>(0.2872) | -0.4284***<br>(0.1152) | -0.4108**<br>(0.1750) |
| *ln(Lung cancer prevalence per 100K)* | 0.6502**<br>(0.2742) | 0.6658*<br>(0.3126) | 0.2843<br>(0.3296) |  |  |  |  | 0.5036<br>(0.3149) | 0.6700**<br>(0.2520) | 0.6647***<br>(0.2520) |
| *Mean body mass index* | -0.0350<br>(0.0433) | -0.0365<br>(0.0449) | 0.1648*<br>(0.0804) |  |  |  |  | 0.2254<br>(0.1168) |  |  |
| *Raised blood pressure prevalence (%)* | 0.0908<br>(0.0587) | 0.0840<br>(0.0699) | 0.1052<br>(0.0888) |  |  |  |  | 0.0852<br>(0.0645) | 0.0900*<br>(0.0400) | 0.0939***<br>(0.0358) |
| *Tuberculosis vaccine coverage (%)* | -0.0035<br>(0.0025) | -0.0053***<br>(0.0014) | -0.0084**<br>(0.0025) |  |  |  |  | -0.0024<br>(0.0029) | -0.0050***<br>(0.0012) | -0.0030<br>(0.0027) |
| **Health Infrastructure Variables** |  |  |  |  |  |  |  |  |  |  |
| *ln(Nurses & midwives per 10K)* | 0.1988<br>(0.1667) | 0.2175<br>(0.1815) | 0.3173<br>(0.2388) |  |  |  | 0.6244**<br>(0.1872) | 0.3105<br>(0.2614) | 0.1983<br>(0.1641) | 0.1975<br>(0.1534) |
| *Health services effective coverage index* | 0.0424***<br>(0.0093) | 0.0382***<br>(0.0094) | 0.0336**<br>(0.0120) |  |  |  | 0.0784***<br>(0.0165) | 0.0467***<br>(0.0126) | 0.0384***<br>(0.0042) | 0.0425***<br>(0.0036) |
| **World Bank Region Dummy Variables** |  |  |  |  |  |  |  |  |  |  |
| *East Asia & Pacific* | -2.9121***<br>(0.4156) | -2.7222***<br>(0.4126) |  |  |  |  |  |  | -2.7276***<br>(0.2219) | -2.7887***<br>(0.1898) |
| *Europe & Central Asia* | -0.9212***<br>(0.2648) | -0.8984**<br>(0.3429) |  |  |  |  |  |  | -0.9777**<br>(0.3203) | -0.9412***<br>(0.2855) |
| *South Asia* | -0.3021<br>(0.2783) | -0.2916<br>(0.2575) |  |  |  |  |  |  | -0.3033<br>(0.3386) | -0.3653<br>(0.3914) |
| *Middle East & North Africa* | -0.5796***<br>(0.1709) | -0.5164**<br>(0.1952) |  |  |  |  |  |  | -0.5724**<br>(0.2037) | -0.6184**<br>(0.2433) |
| *Sub-Saharan Africa* | -1.8408***<br>(0.2977) | -1.4714**<br>(0.5810) |  |  |  |  |  |  | -1.5385**<br>(0.5503) | -1.839**<br>(0.4449) |
| *Constant* | -3.8209<br>(2.8889) | -3.7009<br>(2.6583) | -6.4935<br>(3.3812) | 5.7771***<br>(0.4472) | -8.9249***<br>(2.2835) | -2.7549<br>(1.9304) | -1.2151<br>(0.6648) | -4.6518<br>(3.3343) | -4.9776**<br>(1.8925) | -4.9776***<br>(1.8925) |
| *N* | 142 | 132 | 132 | 142 | 141 | 138 | 139 | 135 | 135 | 142 |
| *R<sup>2</sup>* | 0.7404 | 0.7288 | 0.6455 | 0.1859 | 0.4337 | 0.5265 | 0.4479 | 0.5876 | 0.7378 | 0.7367 |

Notes: Standard errors in parentheses; two-tailed significance levels \*\*\*0.01, \*\*0.05, \*0.10

## ACKNOWLEDGEMENTS

We thank the *Martindale Center for the Study of Private Enterprise* at Lehigh University for supporting the Data for Impact Fellows program, this research, and related database effort. Thanks too to Vice Provost Khanjan Mehta and William Whitney, both of the *Office of Creative Inquiry* at Lehigh University, who organized and offered the Data for Impact Summer Institute in partnership with the Martindale Center and the Institute for Data, Intelligent Systems, and Computation (I-DISC). We also thank Professor Hank Korth for database structuring assistance.

